# HLA allele-specific expression loss in tumors can shorten survival and hinder immunotherapy

**DOI:** 10.1101/2020.09.30.20204875

**Authors:** Ioan Filip, Rose Orenbuch, Junfei Zhao, Gulam Manji, Evangelina López de Maturana, Núria Malats, Kenneth P. Olive, Raul Rabadan

## Abstract

Efficient presentation of aberrant peptide fragments by the human leukocyte antigen class I (HLA-I) genes is necessary for immune detection and killing of cancer cells. Patient HLA-I genotypes are known to impact the efficacy of cancer immunotherapy, and the somatic loss of HLA-I heterozygosity has been established as a factor in immune evasion. While global deregulated expression of HLA-I has been reported in different tumor types, the role of HLA-I allele-specific expression loss – that is, the preferential RNA expression loss of specific HLA-I alleles – has not been fully characterized in cancer. In the present study, we quantified HLA-I allele-specific expression (ASE) across eleven TCGA tumor types using a novel method from input RNA and whole-exome sequencing data. Allele-specific loss in at least one of the three HLA-I genes (ASE loss) was pervasive and associated to worse overall survival across tumor types, including pancreatic adenocarcinomas, prostate carcinomas and glioblastomas, among others. In particular, our analysis shows that detection of neoantigens with binding affinity to the specific HLA-I genes subject to ASE loss was a top prognostic indicator of overall survival. Additionally, we found that ASE loss hindered immunotherapy in retrospective analyses. Together, these results highlight the prevalence of HLA-I ASE loss – a previously uncharacterized phenomenon in cancer – and provide initial evidence of its clinical significance in cancer prognosis and immunotherapy treatment.

## Introduction

Somatic mutations and chromosomal instability drive carcinogenesis and progression. Mutant peptide fragments derived from aberrant proteins can trigger a cytotoxic T-cell response through recognition of neoantigens that differ sufficiently from the normal host peptides^1^. As expression of HLA-I genes is necessary for neoantigen presentation in cancer cells, disruptions in HLA-I expression can have major implications on immune evasion. Meta-analyses of human cancers indicate abnormal or lowered HLA-I expression in particular for non-small cell lung cancer, breast carcinoma, head-neck squamous cell carcinoma, melanoma, as well as bladder, pancreas and prostate tumors, in up to 90% of primary samples^2, 3,4,5.^ Although downregulation of HLA class I can allow tumor cells to escape immune detection by cytotoxic T-cells, complete HLA-I loss makes cells vulnerable to natural killer (NK) antitumor activity as they are no longer able to present self-antigens on the cell surface^6^. The tumor microenvironment, therefore, plays a critical role in immune escape^3^, and it has been suggested that decreased expression of HLA-I, but not complete loss, can allow tumors to escape from both T-cell and NK surveillance^7^. Downregulation of HLA-I is associated with worse prognosis^3,8^, but it is also associated with a decreased metastatic potential^9^.

Patient HLA-I genotypes are known to impact the efficacy of cancer immunotherapy^10,11^ and the loss of HLA-I germline heterozygosity (LOH) – through partial or full chromosome 6 loss or focal deletion of the HLA locus – is a common molecular mechanism driving abnormal HLA-I expression^5^. LOH, traditionally assessed through analysis of microsatellite markers, is frequently observed in many tumor types such as head-neck^12^ and pancreatic cancer^13^. Haplotype-specific copy number inference through computational approaches has enabled LOH assessment from standard next-generation DNA sequencing, showing that LOH occurs in 40% of non-small-cell lung cancers^14^. While a few tools have recently reported HLA allele-specific quantification of mRNA expression^15^ (ASE), even at single-cell resolution^16^, there is currently no gold standard for HLA-I ASE from RNA-seq data. Furthermore, the clinical significance of HLA-I LOH at the level of expression (ASE loss) is poorly understood across cancer subtypes.

In the present study, we aimed to systematically characterize HLA-I ASE loss across tumor types using a novel, accurate allele-specific quantification method (Figure 1a) that builds upon previously established high-resolution HLA genotyping protocols from RNA-seq^17,18^. In light of ubiquitous HLA-I aberrant expression in cancer, we hypothesized that HLA-I ASE loss may constitute a universal immune escape mechanism with significant clinical impact, particularly in the context of immunotherapy. HLA-I ASE in cancer tissue was first quantified using our tool, *arcasHLA-quant*^*19*^, for 3,585 individuals across eleven TCGA molecular tumor subtypes (Figure 1b) of the brain, head and neck, lung, breast, pancreas, kidney, bladder, prostate and the skin. Paired tumor RNA and DNA sequencing was required for this study, in addition to matched normal DNA controls (see Cohort descriptions in Online Methods). As a consistency check, we verified that gene-specific quantification levels obtained with *arcasHLA-quant* by summing minor and major allele expression for each HLA-I gene (HLA-A, HLA-B and HLA-C), were consistent with expression levels as inferred through alternate methods^20^ available on the TCGA portal (Pearson correlation coefficients in the range 0.64-0.94, *p* < 10^−16^ across tumor subtypes, e.g. BLCA, Supplementary Figure 1a). Next, for each HLA-I gene, we inferred the expressed copy number for each genotyped allele (e.g. HLA-A1 and HLA-A2 in the case of an HLA-A heterozygous individual; see HLA-I genotyping and ASE quantification). Compared to HLA-I expression in normal tissue samples from GTEx^21^, TCGA subtypes showed significant differences in the distribution of minor allele frequencies (Supplementary Figure 1b). Thus, we posited that extensive allelic imbalance observed was due to the tumor component in the bulk samples. Using inferred estimates of tumor purity and ploidy^22^, we then defined tumor HLA-I ASE loss for the TCGA cases with detectably high HLA-I expression imbalance in minor-major allele pairs in any of the class I HLA genes (Figure 1a; see Tumor purity and ploidy inference, and Assessment of ASE loss in Online Methods). Our assessment of HLA-I ASE loss was consistent with detection of nonsense HLA mutations in TCGA (Supplementary Table 1; see ASE loss and nonsense HLA-I mutations in Online Methods).

**Fig. 1.**
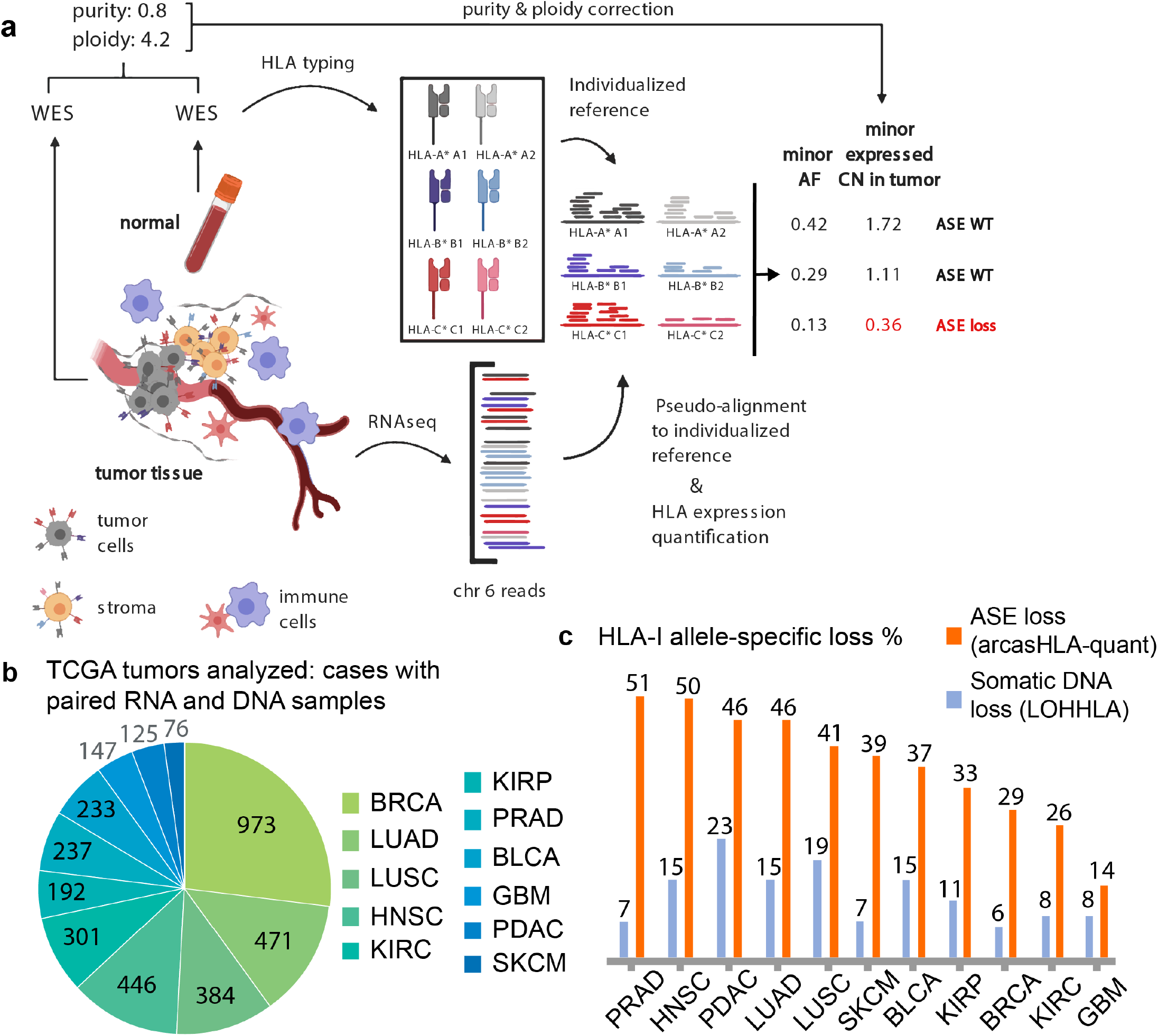
HLA class I allele-specific expression loss is pervasive in cancer. **a**. Workflow for assessment of HLA-I allele-specific expression loss (ASE loss) in bulk RNA-seq, from input RNA and whole-exome (WES) sequencing data. **b**. TCGA subtypes analyzed in this work (project names listed clock-wise in the pie chart). **c**. Proportion of HLA-I ASE loss across TCGA subtypes (orange) as inferred using *arcasHLA-quant*, and proportion of cases where expression loss is accompanied by somatic DNA loss (blue), as inferred with the LOHHLA tool from WES data. (Panel a. was created with BioRender.)

## Results

### HLA-I allele-specific expression loss is pervasive across tumor types

We first determined that HLA-I ASE loss is pervasive across TCGA tumor subtypes^23^ (Figure 1b, c): ASE loss was detected in every tumor type analyzed, with prostate (PRAD), head and neck (HNSC), pancreatic (PDAC) and lung adenocarcinomas (LUAD) exhibiting HLA-I ASE loss at frequencies above 45% in their respective cohorts, while glioblastomas (GBM) showed a markedly lower incidence of ASE loss at 14%. Overall, HLA-I ASE loss was due to HLA-A in 55% of cases, to HLA-B in 39% and to HLA-C in 45% of cases, with loss at all three genes occurring at a rate of 23%. We then asked whether ASE loss was accompanied by somatic DNA lesions (e.g. chromosomal or focal deletions) at the corresponding HLA-I loci. Using LOHHLA^14^, we found that only a fraction of ASE losses showed evidence of DNA haplotype loss (Figure 1c; see Assessment of somatic loss of HLA-I haplotypes). The maximal proportion of DNA-to-expression-only loss was found in TCGA-GBM (57%), while TCGA-PRAD had the smallest such proportion (under 14%). Our estimates of ASE loss are consistent with established literature on several tumor types. For example, in pancreatic cancer (TCGA-PDAC), previous studies^13^ indicate that LOH occurs in 21% of cases, while expression loss without LOH occurs much more frequently (in 58% of cases). Other reports find higher incidence of somatic LOH than what we show here. In head-neck cancer (TCGA-HNSC) for instance, LOH was previously detected in 49% of cases with deregulated HLA-I expression^12^ (in our data, 30%). More notably, in non-small-cell lung cancer, LOH was detected in 40% of cases^14^, while in our data we found somatic loss in about 15-19% of cases (Figure 1c). These discrepancies may possibly be due to differences in cohort selections, methodologies, sequencing depth, or smaller sample sizes in previously published assessments of LOH and HLA-I expression. Nonetheless, our results suggest that the majority of HLA-I ASE loss occurs in cancer occurs through epigenetic or other expression regulatory mechanisms instead of somatic DNA lesions.

### HLA-I allele-specific expression loss decreases overall survival in cancer

Next, we hypothesized that presence of HLA-I ASE loss might lead to shorter survival times across tumor types, owing to a gain of immune escape potential. In order to conduct a pan-cancer analysis of the clinical significance of ASE loss, we defined thirty tumor sample features to be incorporated into a multivariable Cox regression stratified by tumor type (Supplementary Table 2). In addition to age at diagnosis, tumor purity and ploidy estimates, we focused on immune-related and microenvironmental features. To that end, we included HLA-I patient genotype (at the level of HLA allele supertypes^24^ and HLA-I germline homozygosity; see HLA-I genotyping supertypes in Online Methods), and immune cell subtype proportions as inferred through *in silico* decomposition methods^25^ (including CD4+ and CD8+ T-cells, B-cells, NK-cells and macrophages; see *In silico* decomposition into immune cell subtypes). Finally, we included several features related to predicted neoantigens, HLA allele-specific neoantigen affinities and mutational burden in the tumor samples^26^ (see Computational identification of neopeptides). Our Cox proportional-hazards model of overall survival (n = 3386, with 2453 censored events), stratified across all eleven TCGA molecular subtypes and including all features, showed a trend for HLA-I ASE loss contribution towards worse prognosis (*p* = 0.07; hazard ratio, HR, = 1.17; 95% confidence interval, CI: 0.99-1.37; Supplementary Table 2). However, the largest-effect predictor of poorer overall survival was the neoantigen count adjusted for ASE status of the HLA-I allele with the highest binding affinity (*p* = 0.01, HR = 1.38, 95% CI: 1.08-1.77; or HR = 1.02 per neoepitope; see Figure 2a, Supplementary Figure 2, Supplementary Table 2). Consistent with this observation, we found that detection of neoantigens with binding affinity to kept alleles had an overall beneficial effect in the pan-cancer analysis (*p* = 0.03, HR = 0.83, 95% CI: 0.7-0.98; or HR = 0.98/neoepitope). Predictive features based on neoantigen counts with major and minor HLA-I allele affinities respectively, independent of ASE loss, did not have the same significant effects on overall survival (Supplementary Table 2). Two other variables were associated with shorter overall survival: age at diagnosis (*p* < 0.005, HR = 1.31, 95% CI: 1.21-1.40; or HR = 1.02/year), and tumor ploidy (*p* < 0.005, HR = 1.16, 95% CI: 1.08-1.25). ASE loss showed a trend towards shortening survival in seven tumor subtypes: PRAD, HNSC, PDAC, LUSC, KIRP, KIRC and GBM (Supplementary Tables 3, 4a; see Statistical analyses); but our ASE data were not significant in separate analyses by tumor subtype – due to a lack of power, differences in the quality and quantity of neoantigen predictions across TCGA, and gaps in characterizing the phenotypes of tumor-infiltrating immune cells (such as T-cell exhaustion^27^). In the high-powered pan-cancer analysis, however, our data suggest a significant clinical impact of HLA-I ASE loss towards poor prognosis, particularly when the loss phenotype is accompanied by an increased count of neoantigens predicted to have a higher binding affinity to the alleles that are lost.

**Fig. 2.**
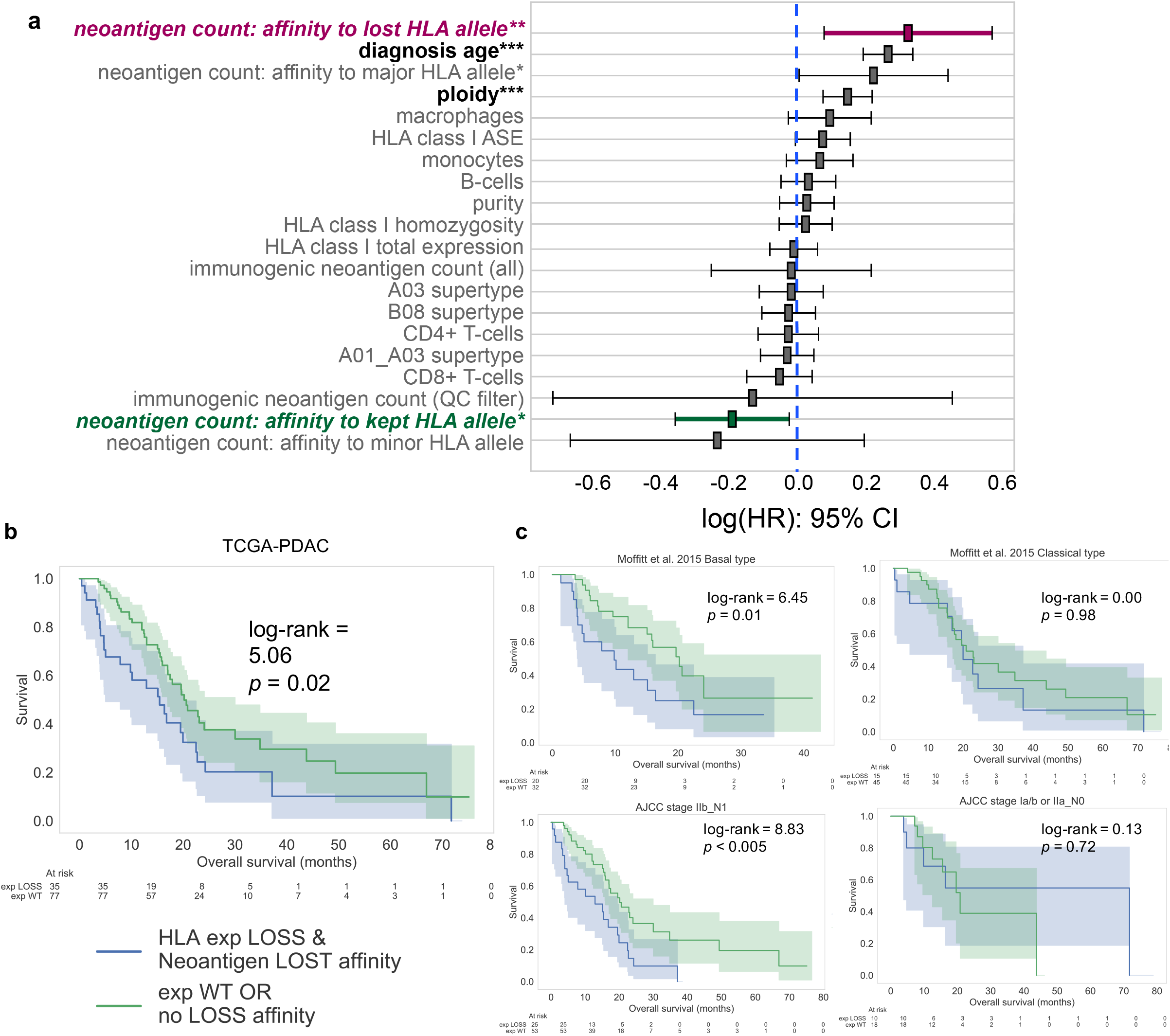
HLA class I allele-specific expression loss contributes to worse overall survival in cancer. **a**. Prognostic features in a pan-cancer Cox proportional-hazards model of overall survival, stratified by tumor type. Thirty features were standardized (mean 0, standard deviation 1) and included (full list in Supplementary Figure 2 and Table 2). We incorporated neoantigen levels accounting for ASE status of HLA-I alleles with the highest predicted binding affinity, and counted neoantigens with higher affinity towards either the “kept” or the “lost” alleles (if any). The neoantigen count with lost-allele affinity was the largest-effect predictor of survival. **b**. Survival curves in TCGA-PDAC: cohort with HLA-I ASE loss and simultaneous detection of neoantigens with predicted affinity towards genes subject to loss has a worse prognosis. **c**. Survival curves of TCGA-PDAC according to transcriptional subtype and tumor stage.

### HLA-I allele-specific expression loss contributes to decreased overall survival in PDAC

Next, in order to mitigate potential limitations of modeling allele-specific binding, we only considered neoantigen counts adjusted for the ASE status of the HLA-I gene with the highest predicted affinity. In addition, since tumor purity was associated with ASE loss (Supplementary Table 5), in order to further limit ASE loss calling errors due to extremely low purity levels, we also filtered out TCGA samples with purity below 10% in subsequent revised survival analyses (see Cohort descriptions). In these revised subtype-specific analyses (Supplementary Table 4b; Supplementary Figure 3), a single cancer type showed a significant association with overall survival: TCGA-PDAC^28^ (*p* = 0.03, HR = 1.30, 95% CI: 1.03-1.63; or HR = 1.75/neoepitope). Specifically, the subgroup with worse prognosis exhibited HLA-I ASE loss concomitant with detection of neoantigens predicted with higher affinity towards HLA-I genes subject to expression loss (Figure 2b). Even without accounting for neoantigen HLA-I affinities, ASE loss alone contributed to worse survival in PDAC (*p* = 0.23, HR = 1.34, 95% CI: 0.84-2.20; Supplementary Figure 4).

Next, we validated widespread HLA-I ASE loss in an independent cohort of 96 laser-capture micro-dissected pancreatic ductal adenocarcinoma samples^29^ where RNA-seq was performed separately on cleanly delineated epithelial and stroma compartments (CUMC cohort: CUMC-E for epithelial samples and CUMC-S for the stroma; see Cohort descriptions in Online Methods). Indeed, HLA-I ASE loss was strongly associated with the tumor epithelial compartment (Fisher exact test, OR = 3.98, *p* < 10^−5^), which further supports our hypothesis that ASE loss occurs in the cancer cells from bulk mixtures. Additionally, HLA-I ASE loss resulted in shorter survival when detected in CUMC-E (*p*-val = 0.05, HR = 1.59, 95% CI: 1-2.51). There was no survival impact, however, for ASE loss in CUMC-S (Supplementary Figure 5).

We subsequently interrogated potential associations between HLA-I ASE loss in pancreatic cancer and well-characterized transcriptional subtypes (classical versus basal)^30,31,32,33^, as well as tumor stage. We found that ASE loss in PDAC was present in both subtypes from TCGA, but that it was enriched in basal tumors (OR = 1.88, *p* = 0.15). Moreover, detection of HLA-I ASE loss in the basal subtype, but not in the classical one, was associated with worse survival, suggesting the existence of a basal-like subcategory of PDAC characterized by HLA-I ASE loss and poorer prognosis (Figure 2c). These results were consistent with alternate definitions of transcriptional subtypes (Supplementary Figures 6, 7). Finally, ASE loss detected at AJCC stage 2B had a significant impact towards shorter survival; ASE loss was detected in earlier stages too, although without noticeable clinical effect (Figure 2c). The association to transcriptional subtype was also validated in CUMC-E, where HLA-I ASE loss was detected predominantly in tumors with squamous features (OR = 4.02, *p* = 0.11). Altogether, our findings indicate that HLA-I allele-specific expression loss is a prognostic marker of shorter overall survival in pancreatic ductal adenocarcinoma, particularly in squamous-type, later stage tumors.

### HLA-I allele-specific expression loss decreases efficacy of anti-PD-1 immunotherapy in metastatic melanoma

Finally, we hypothesized that HLA-I ASE loss could be a factor in the efficiency of immune checkpoint blockade immunotherapies. We revisited a previously published metastatic melanoma cohort^34^ with pre-treatment (n = 46) and on-treatment (n = 29) samples (see Cohort descriptions) and inferred HLA-I ASE loss as described before. As in TCGA, HLA-I ASE loss was largely due to minor-major allelic imbalance in the bulk samples (Supplementary Figure 8). Excluding samples with ultra-low tumor purity (purity below 10%), we found ASE loss both pre- and on-Nivolumab therapy in about 37% of the cases (Figure 3). Furthermore, ASE loss contributed to worse overall survival whether assessed before or during therapy. The group with on-therapy ASE loss showed a slightly greater effect on prognosis (*p* = 0.10, HR = 2.28, 95% CI: 0.85-6.11). It is well known that HLA class I homozygosity can reduce overall survival with immune checkpoint blockade^10^ (Supplementary Figure 9). As such, we also analyzed the impact of ASE loss separately for individuals heterozygous at all three HLA-I genes (henceforth “fully heterozygous” cohort). For these individuals, ASE loss resulted in significantly worse prognosis (*p* = 0.02, HR = 4.28, 95% CI: 1.22 – 15.0; Figure 3) when expression loss occurred on-therapy (that is, one-month after the start of therapy^34^). To a large extent, decreased survival might be predictable for heterozygous individuals even before therapy, although the results are not as conclusive pre-treatment (*p* = 0.1, HR = 2.15, 95% CI: 0.85-5.42). By taking neoantigen predictions into account, the survival impact was more striking for the fully heterozygous cohort (*p* = 0.02, HR = 6.24, 95% CI: 1.34-29.1; Supplementary Figure 10). Results with the full cohort (including cases with ultra-low tumor purity) showed the same trend towards worse prognosis, particularly for fully heterozygous individuals with on-treatment ASE loss (*p* = 0.06, HR = 3.25, 95% CI: 0.97-10.9; Supplementary Figure 11). Interestingly, among the fully heterozygous individuals with on-treatment samples and RECIST v1.1-defined response (n = 17), there were only 3 responders (complete or partial), and all of them had the HLA-I ASE wildtype phenotype (OR = inf., *p* = 0.21). Among pre-treatment samples, ASE loss resulted in suggestively lower odds of responding to subsequent treatment (OR = 0.67, n. s.). In addition, survival associations were not explained by factors such as sample purity or somatic LOH (Supplementary Figures 12, 13). Therefore, our results highlight a potential significant clinical impact of HLA-I ASE loss on the efficacy of anti-PD-1 immunotherapy in metastatic melanoma.

**Fig. 3.**
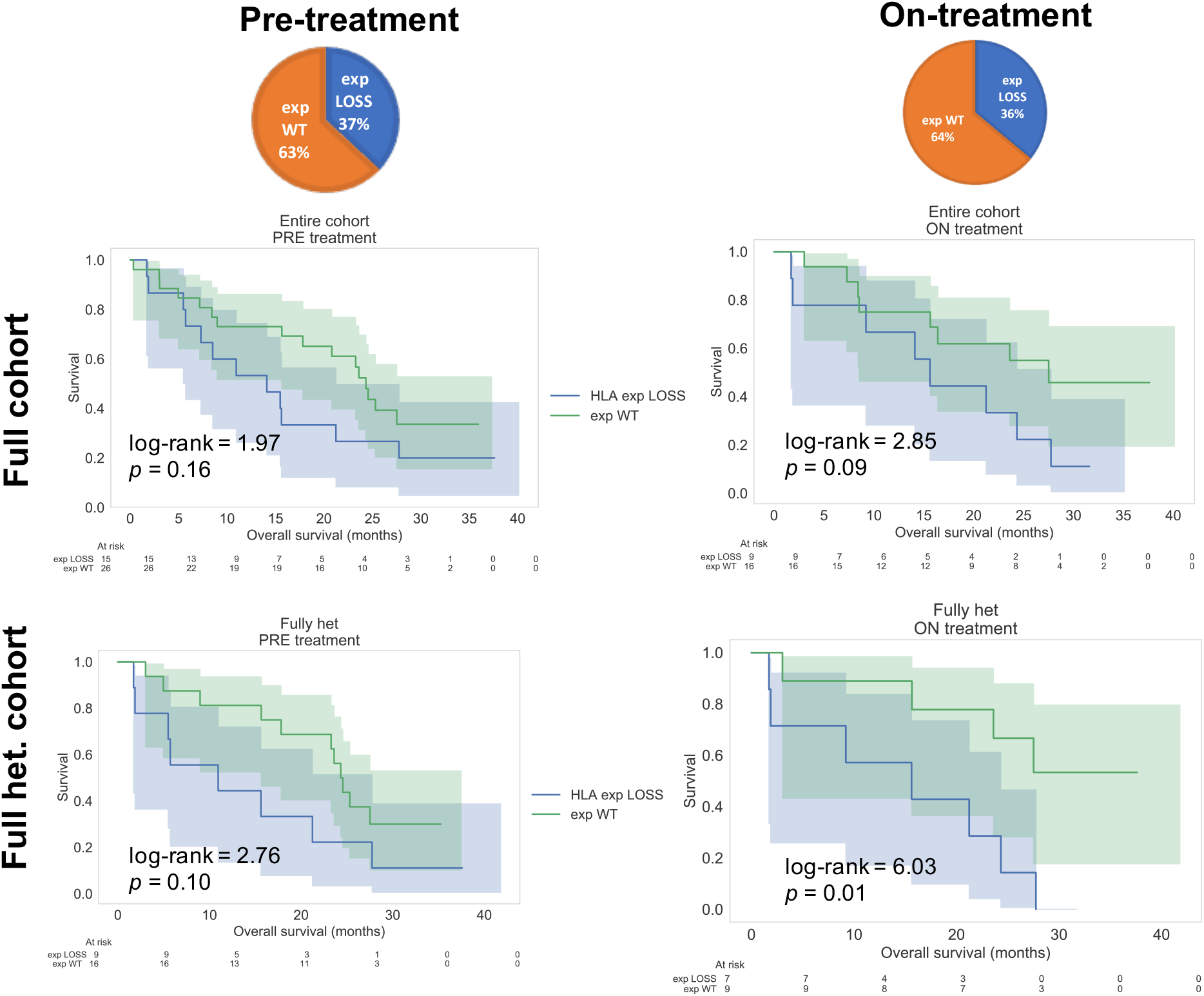
HLA class I allele-specific expression loss decreases efficacy of anti-PD-1 immunotherapy in metastatic melanoma cohort (Riaz *et al*., 2017). Extensive HLA-I ASE loss in melanoma cohort pre- and on-treatment with Nivolumab. Fully heterozygous individuals with ASE loss detected on-treatment show significantly worse overall survival.

## Conclusion

Altogether, our work demonstrates that HLA-I ASE loss – as inferred through our allele-specific RNA-seq quantification method, *arcasHLA-quant* – is a frequent phenomenon across tumor types. Moreover, the majority of ASE loss cases do not seem to result from somatic DNA deletions in HLA-I. In a pan-cancer analysis, among predictor variables related to HLA-I and immune cell infiltration, we found that detection of neoantigens with binding affinity to specific class I genes subject to ASE loss was a top prognostic indicator of overall survival. In addition, ASE loss was shown to contribute to a worse prognosis in PDAC, as well as in metastatic melanoma under immune checkpoint blockade therapy. Our study has some limitations related to the lack statistical power in stratified analyses, the difficulty of HLA allele-specific quantification, the challenges of computational neoantigen binding prediction and the complexity of overall survival outcomes. However, the prevalence of ASE loss and the initial clinical impact that we have established here should highlight the importance of further investigations of HLA-I ASE in cancer, with the goal of understanding the underlying mechanisms and the timing of this potentially reversible lesion in tumor evolution.

## Data Availability

Data is in public data repositories.

## Author Contributions

I.F. and R.R. designed the study. I.F. and R.O. designed the HLA quantification algorithm. K.P.O. conducted the CUMC cohort study and G.M reviewed the clinical annotation for CUMC. I.F., J.Z. and R.O performed data processing and computational analyses. R.R. and N.M. co-led the PDAC analyses. All authors contributed to the manuscript.

## Acknowledgements

We gratefully acknowledge the Pancreatic Cancer Collective Grant from SU2C and the Lustgarten Foundation, as well as funding from NCI/NIH grant U01CA243073. We also acknowledge the guidance of Anil K. Rustgi on this project. The CUMC cohort samples were procured by the HICCC Molecular Pathology Share Resource with support from, and under the guidance of Hanina Hibshoosh. Outcome data for the CUMC cohort were curated by the HICCC Database Shared Resource under the guidance of Jeanine Genkinger. We also thank Karen Gomez for helping us process and analyze TCGA data.

## Disclosure of Potential Conflicts of Interest

R.R. is a founder and a member of the SAB of Genotwin, and a member of the SAB of AimedBio.

## Online Methods

### Cohort descriptions

#### TCGA

We have included 3585 tumors from TCGA^23^ across eleven molecular subtypes (breakdown illustrated in Figure 1b) where whole exome sequencing (WES) samples were available from the tumor and from normal tissue, in addition to matched RNA-seq derived from the same tumor sample. ASE loss ratios were calculated (Figure 1c) based on these TCGA cases. For survival analyses and other clinical associations, we excluded cases without age at diagnosis information and cases without overall survival data. In total, 3386 cases were included in the Cox regression analyses (Figure 2a). For the TCGA-PDAC dataset, we eliminated TCGA cases that were not classified as pancreatic ductal adenocarcinoma using the criteria laid out in the following reference: Nicolle, R. *et al*., 2019. In addition, we excluded samples with ultra-low purity estimate (sequenza-inferred purity below 10%; see Tumor purity and ploidy inference below) in subsequent survival analyses. In total, 3187 cases satisfied the purity condition (purity > 0.1). The TCGA-PDAC cohort consisted of 122 individuals with DNA/RNA data, age and overall survival information; and 112 individuals with the added tumor purity criterion.

#### GTEx

We also included an arbitrary selection of samples from GTEx^21^ from bladder (all samples available for download, n = 11), brain (n = 21), lung (n = 21), pancreas (n = 21) and skin tissues (n = 21). These samples served as normal controls for HLA-I allele-specific expression imbalance levels (Supplementary Figure 1b).

#### CUMC cohort

We analyzed a previously published cohort at Columbia University^29^ (denoted as CUMC, n = 192) comprised of epithelial samples (CUMC-E, n = 96) and stroma samples (CUMC-S, n = 96) that were cleanly delineated through laser-capture microdissection and subsequently processed and sequenced separately. All n = 96 cases were diagnosed as PDAC, and had overall survival information available. The vast majority of samples (94%) were stage 2A or 2B. **Riaz *et al***., **2017**. For the analysis of ASE loss in metastasis, we included a retrospective study of pre- and on-treatment samples in metastatic melanoma^34^. In all, we identified n = 75 cases with paired DNA and RNA samples as required for our pipeline (Figure 1a). For all analyses with the Riaz dataset (Figure 3), we excluded cases with ultra-low purity (sequenza-inferred purity below 10%): there were n = 41 pre- and n = 25 on-treatment cases remaining.

### HLA-I genotyping and HLA supertypes

For all the TCGA cohorts in this study, high-resolution HLA class I genotyping (performed with Polysolver^35^ from normal WES samples) was previously available^23^. For the metastatic melanoma cohort, the HLA-I genotypes were also previously available^10^. For the CUMC and the GTEx cohorts, high-resolution HLA-I genotyping was performed from RNA-seq using arcasHLA^17^. In the former, only the stromal compartment was used to infer patient HLA genotypes. All HLA supertypes^24^ were annotated in the TCGA cohort for each subject and included as binary predictor variables in the multivariate Cox regressions.

### HLA-I allele-specific expression (ASE) quantification

*arcasHLA-quant*^19^ quantifies allele-specific and gene-level expression given an individual’s genotype, that is either determined using *arcasHLA* from RNA-seq (as for the CUMC cohort in pancreatic cancer) or through DNA-based methods (eg. Polysolver) when normal WES samples are available. Similar to existing approaches^15^, the *arcasHLA-quant* method first builds a customized transcriptome reference by replacing the default HLA transcripts from the human chromosome 6 reference (GRCh 38) with patient-specific HLA-I allelic cDNA references obtained from the IMGT/HLA database^36^. Subsequently, reads from input BAM files are extracted as in *arcasHLA*, and allele-specific expression quantification is performed using Kallisto^37^. This approach extends the workflow and applicability of *arcasHLA*; importantly, the same pipeline for extracting reads from input samples and constructing graph-based references for pseudo-mapping – which give *arcasHLA* high-resolution accuracy in genotyping HLA class I and class II genes from RNA-seq – are used for *arcasHLA-quant*. The *arcasHLA-quant* pipeline is developed in Python and can be run as a command-line instruction set or in a virtual environment. It is publically available: https://github.com/roseorenbuch/arcasHLA-quant.

### Tumor purity and ploidy inference

We used the sequenza^22^ algorithm with default parameters to obtain purity and ploidy estimates for all the TCGA samples, and likewise for the samples from the metastatic melanoma cohort (Riaz *et al*., 2017). Among all the solutions proposed by the model, we selected the purity-ploidy pair with the highest posterior probability. For the purpose of calculating ASE loss in the CUMC cohort, because we did not have DNA sequencing available, we assumed that the laser-capture microdissected CUMC-E and CUMC-S samples had 100% purity, and ploidy equal to 2.0. For comparison, the mean TCGA-PDAC purity was 53%, with a standard error of the mean (SEM) equal to 2.6%, while the mean ploidy was 2.02 with SEM equal to 0.05.

### Assessment of allele-specific expression loss (ASE loss)

In order to determine the status of HLA-I ASE loss in the tumor component of bulk RNA-seq, we incorporated the following two pieces of information: tumor purity and ploidy inferred from paired tumor and normal samples; and HLA ASE inferred from RNA-seq using *arcasHLA-quant*. Similar to a previously published criterion for somatic LOH, LOHHLA^14^, we first determined a purity- and ploidy-adjusted tumor expressed copy number (exp_CN_) for each HLA-I allele, as follows:

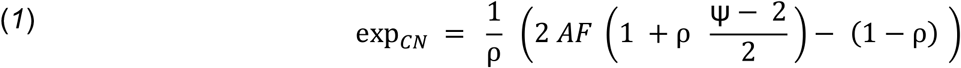

where *AF* denotes the allelic frequency (namely, the ratio of reads attributed to each allele over the total read count for the corresponding HLA gene) in the bulk sample, ρ denotes the tumor purity and Ψ, the overall tumor ploidy (obtained from sequenza^22^). Second, as in LOHHLA, we defined ASE loss as the occurrence of a minor allele exp_CN_ below 0.5 in at least one HLA-I gene (HLA-A, -B or -C). We note that formula (*1*) yields an expressed copy number (exp_CN_) of 1 for both minor and major alleles in the case of a heterozygous class I HLA gene with perfectly balanced allelic expression levels (i.e. with *AF* = 0.5), and 100% tumor purity (ρ = 1.0) and normal ploidy (Ψ = 2.0).

### ASE loss and nonsense HLA-I mutations

Disrupted HLA-I expression can also result from the accumulation of somatic mutations in HLA-I^5^. However, somatic mutations in HLA-I are relatively infrequent in TCGA, varying from below 1% incidence in BRCA and GBM, to around 5% in BLCA, LUAD and SKCM, and up to 10% in HNSC^8,35^. Using a comprehensive list of HLA-I mutations in TCGA^38^, we identified 22 nonsense HLA-I mutations in our cohort in the following subtypes (Supplementary Table 1): HNSC (15), LUSC (2), BLCA (2), LUAD (1), KIRP (1), SKCM (1). We found that 10/22 cases with nonsense mutations were annotated as ASE loss at the correct HLA-I gene, with an additional set of 6/22 cases exhibiting a less stringent measure of allelic imbalance (specifically, minor-to-major allelic ratio < 0.5) but no ASE loss as we have defined it. Only 5/22 cases with nonsense HLA-I mutations showed no allelic expression imbalance in our analysis (in one TCGA-HNSC case, an HLA-A homozygous individual had an HLA-A nonsense mutation where ASE loss could not be assessed). Therefore, our quantification approach is consistent with detection of nonsense HLA mutations in 76% (16/21) of TCGA cases, suggesting enough sensitivity suitable for capturing putative nonsense-mediated decay in bulk RNA-seq^39^.

### Assessment of somatic loss of HLA-I haplotypes

We used LOHHLA^14^ to infer HLA-I allele-specific copy number variation and determine somatic LOH at the level of DNA, from input tumor and normal paired WES samples (see Figure 1c). We set the minimum coverage threshold at 5, and used the default configuration for all other parameters. In this study, we focused on somatic LOH cases that also exhibited HLA-I ASE loss (Figure 1c). We used the criteria for LOH-positive as indicated by LOHHLA, namely: allelic copy number (CN) < 0.5 and *p*-value < 0.05. As such, this led to the inclusion of all cases with bi-allelic DNA loss and with bi-allelic negative CNs within the LOH-positive cases. Cases that resulted in LOHHLA errors were excluded from the comparison with *arcasHLA-quant* ASE loss.

### *In silico* decomposition into immune cell subtypes

We used the CIBERSORT^25^ LM22 signature matrix containing twenty-two functionally defined human immune-cell subtypes in order to quantify the immune cell infiltration in the tumor RNA-seq samples. We used the CIBERSORT support-vector machine approach with default parameters for each sample in TCGA. However, since the method produces a weight decomposition of each bulk sample into fractional contributions from each immune subtype that sum to 1, this method is not entirely adequate for separating tumor cell signatures from immune cell signatures since it does not include a tumor component in the final decomposition. Owing to a lack of normal tissue expression signatures for each corresponding TCGA cohort in our study, for each tumor bulk sample we corrected every immune cell proportions by only retaining the immune cell subtypes reported by CIBERSORT that had a fractional contribution exceeding 10%. Subsequently, we defined the following immune features for Cox regression analyses (Figure 2a) by adding the latter corrected LM22 subtype fractional parts according to their corresponding immune lineage category: CD4+ T-cells, CD8+ T-cells, B-cells, Macrophages, NK-cells and Other Macrophages. For example, the B-cell category was defined as the sum of corrected proportions for the following LM22 subtypes: “B cells memory”, “B cells naïve” and “Plasma cells”. Among the corrected proportions of tumor-infiltrating immune cell types in the full-scale Cox regression analysis, we found that macrophages contributed to worse prognosis (HR = 1.10; not significant, n.s.), while CD4+ and CD8+ T-cells decreased the hazard rate of death (HRs = 0.97, 0.95 resp.; n. s.). Both of these indications, although not statistically significant in this study, are consistent with existing literature that highlights an unfavorable role for tumor-associated macrophages^40^ and a favorable effect of cytotoxic T-lymphocytes targeting cancer cells^41^.

### Computational identification of neopeptides

We used the pVAC-seq pipeline^26^ with the NetMHCcons binding strength predictor to identify neoantigens^42^. NetMHCcons integrates three state-of-the-art methods NetMHC, NetMHCpan and PickPocket to give the most accurate predictions^42^. As required, we used the variant effect predictor from Ensembl to annotate variants for downstream processing by pVAC-Seq^43^. For each single-residue missense alteration, HLA-I allele-specific binding affinities were predicted for all the wild-type and mutant peptide fragments of varying lengths (from 8 to 11 amino acids). The mutant peptide with the strongest binding affinity was kept for downstream analysis. The total immunogenic neoantigen count (Figure 2a) was determined for each individual as the number of predicted mutant epitopes with a median IC50 score below 500. This feature, called “immunogenic neoantigen count (all)”, was subsequently included in the Cox model of overall survival. And additional count of immunogenic antigens was determined after applying the following conservative quality filter: tumor RNA variant allele frequency (VAF) > 0.1, tumor DNA VAF > 0.1, tumor RNA depth > 0, tumor DNA depth > 2 and (normalized) gene expression > 1. The filtered neoantigen count predictor variable was called “immunogenic neoantigen count (QC filter)”. Only total immunogenic neoantigens were included in the variables adjusted for HLA-I ASE loss status, “neoantigen count: affinity to lost HLA allele” and “neoantigen count: affinity to kept HLA allele”; as well as those adjusted for ASE status: “neoantigen count: affinity to minor HLA allele” and “neoantigen count: affinity to major HLA allele”.

### Statistical analyses

Comparisons between total HLA-I expression by HLA gene, as inferred through *arcas-quant* and through alternate methods available on the TCGA portal, were calculated using the Pearson correlation (Supplementary Figure 1a). Comparisons between minor allele frequencies of HLA-I alleles across eleven TCGA subtypes and GTEx normal samples were assessed with the Mann-Whitney and Kolmogorov-Smirnov 2-sample tests (Supplementary Figure 1b). For all multivariable survival analyses and tumor subtype stratification, we used the Cox proportional-hazards model (Figure 2a). HRs for each of the thirty tumor features included in Figure 2a (and Supplementary Figure 2, Supplementary Table 2) were reported after feature standardization (to mean 0 and standard deviation 1). For binary variables, HRs were also reported without standardization. *P*-values for the HRs were reported for the two-tailed z-test of each coefficient in the Cox regression. For the risk assessment of ASE loss in different cohorts, Kaplan-Meier curves were also plotted and *p*-values of the log-rank test were reported (Figures 2b and 3; Supplementary Figures 3 – 7, 9 – 13). The full-scale Cox regression model including all thirty immune and HLA-related predictor variables was underpowered when deployed separately to each TCGA subtype (only TCGA-SKCM gave a significant likelihood ratio test < 0.05 with Bonferroni correction; Supplementary Table 3). Instead, for each molecular subtype in TCGA we analyzed univariate Cox regression models, as well as a simplified model using only 6 predictor features: age at diagnosis, ploidy, HLA-I ASE loss, as well as macrophage, CD4+ and CD8+ T-cell proportions (Supplementary Table 4a). ASE loss was associated with worse prognosis in 7/11 tumor subtypes (PRAD, HNSC, PDAC, LUSC, KIRP, KIRC and GBM), it had no survival effect in LUAD, and was seen to contribute, paradoxically, towards longer survival in 3/11 subtypes (SKCM, BLCA and BRCA); however, with the exception of GBM, none of these associations with HLA-I ASE loss were significant. At the same time, in univariate regressions, the neoantigen count with binding specificity towards lost HLA-I alleles contributed towards shorter overall survival in 6/11 tumors, while the neoantigen count with HLA-I kept-allele affinity also showed improved survival in 6/11 (non-significant results). For analyses of transcriptional subtype associations with ASE loss in pancreatic ductal adenocarcinoma, for analyses of clinical response data in the CUMC cohort, and for enrichments of ASE loss in CUMC-E as compared to CUMC-S, *p*-values and ORs were calculated with the two-tailed Fisher exact test. All correlation, comparative and survival analyses were performed in Python (version 2.7) using the following packages: numpy (version 1.15.4), scipy (version 1.0.1) and lifelines (version 0.17.0).

## Notes

### Competing Interest Statement

Raul Rabadan is a founder and a member of the SAB of Genotwin, and a member of the SAB of AimedBio.

### Funding Statement

Pancreatic Cancer Collective Grant (SU2C/Lustgarten Foundation)
and NCI/NIH grant U01CA243073

### Author Declarations

No specific IRB protocol as data can be found in public repositories.

