## Supplementary Figures for "HLA allele-specific expression loss in tumors can shorten survival and hinder immunotherapy"

### Slide 1
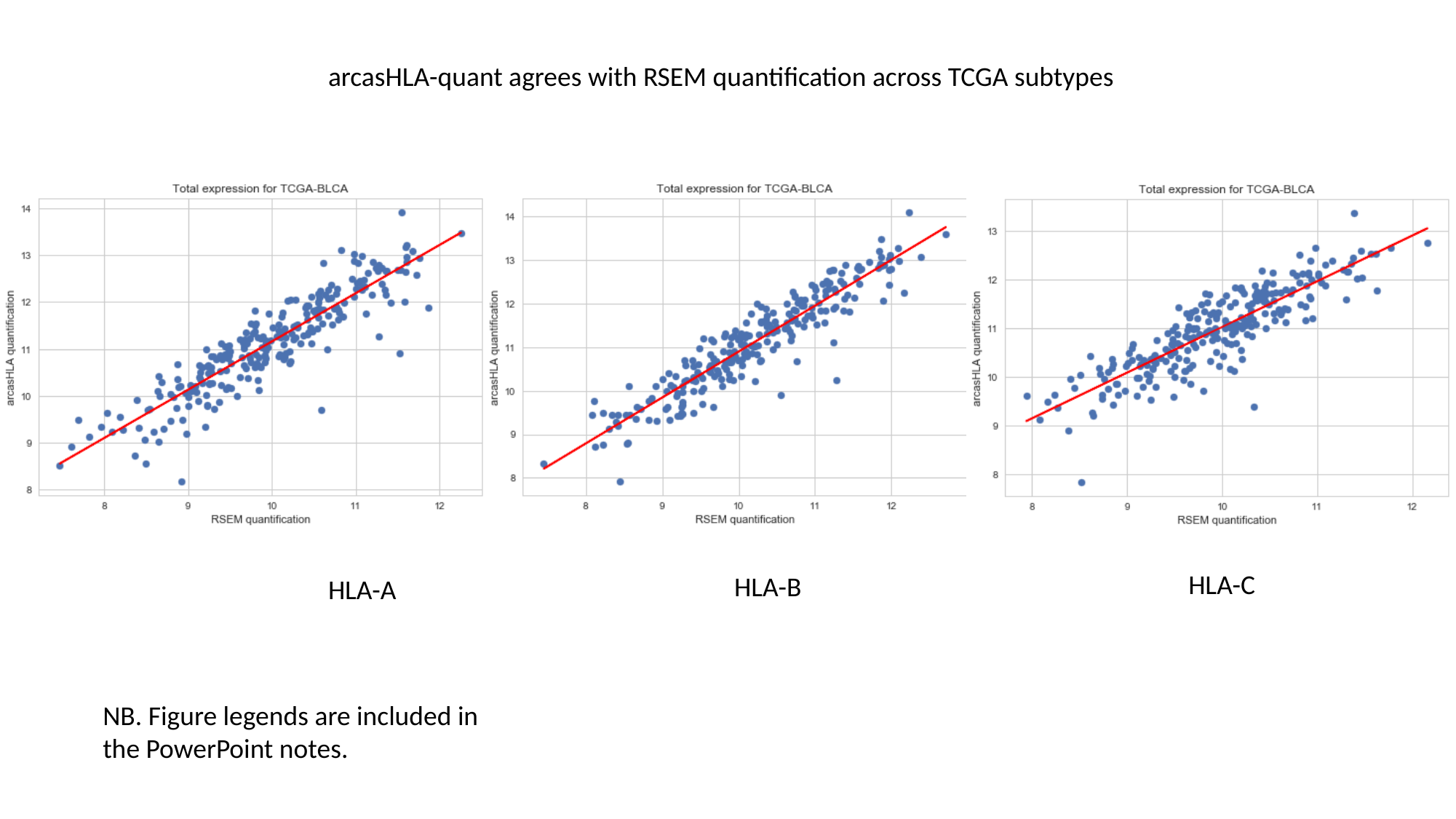

arcasHLA-quant agrees with RSEM quantification across TCGA subtypes
HLA-C
HLA-B
HLA-A
NB. Figure legends are included in the PowerPoint notes.

### Slide 2
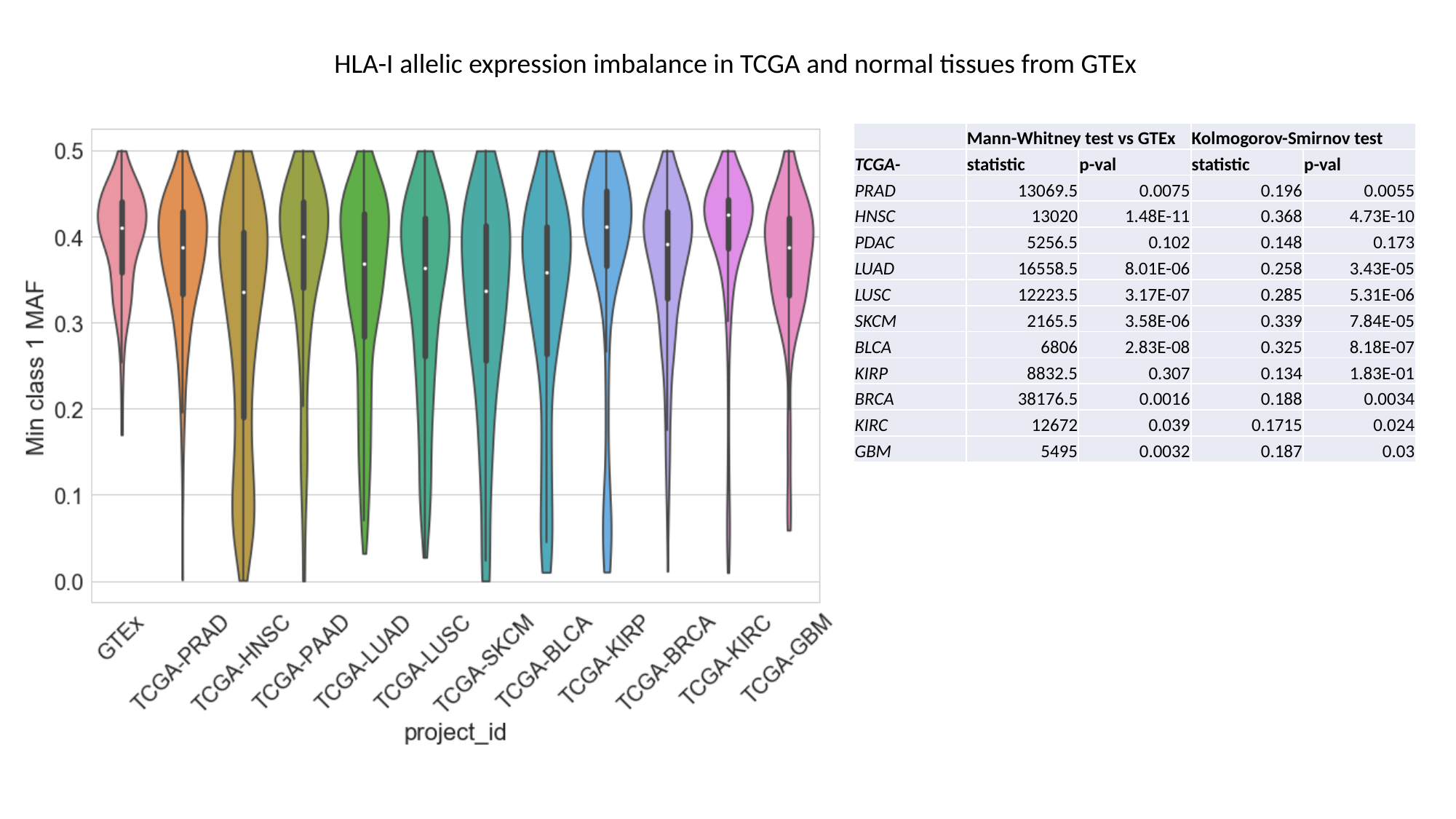

HLA-I allelic expression imbalance in TCGA and normal tissues from GTEx
| | Mann-Whitney test vs GTEx | | Kolmogorov-Smirnov test | |
| --- | --- | --- | --- | --- |
| TCGA- | statistic | p-val | statistic | p-val |
| PRAD | 13069.5 | 0.0075 | 0.196 | 0.0055 |
| HNSC | 13020 | 1.48E-11 | 0.368 | 4.73E-10 |
| PDAC | 5256.5 | 0.102 | 0.148 | 0.173 |
| LUAD | 16558.5 | 8.01E-06 | 0.258 | 3.43E-05 |
| LUSC | 12223.5 | 3.17E-07 | 0.285 | 5.31E-06 |
| SKCM | 2165.5 | 3.58E-06 | 0.339 | 7.84E-05 |
| BLCA | 6806 | 2.83E-08 | 0.325 | 8.18E-07 |
| KIRP | 8832.5 | 0.307 | 0.134 | 1.83E-01 |
| BRCA | 38176.5 | 0.0016 | 0.188 | 0.0034 |
| KIRC | 12672 | 0.039 | 0.1715 | 0.024 |
| GBM | 5495 | 0.0032 | 0.187 | 0.03 |

### Slide 3
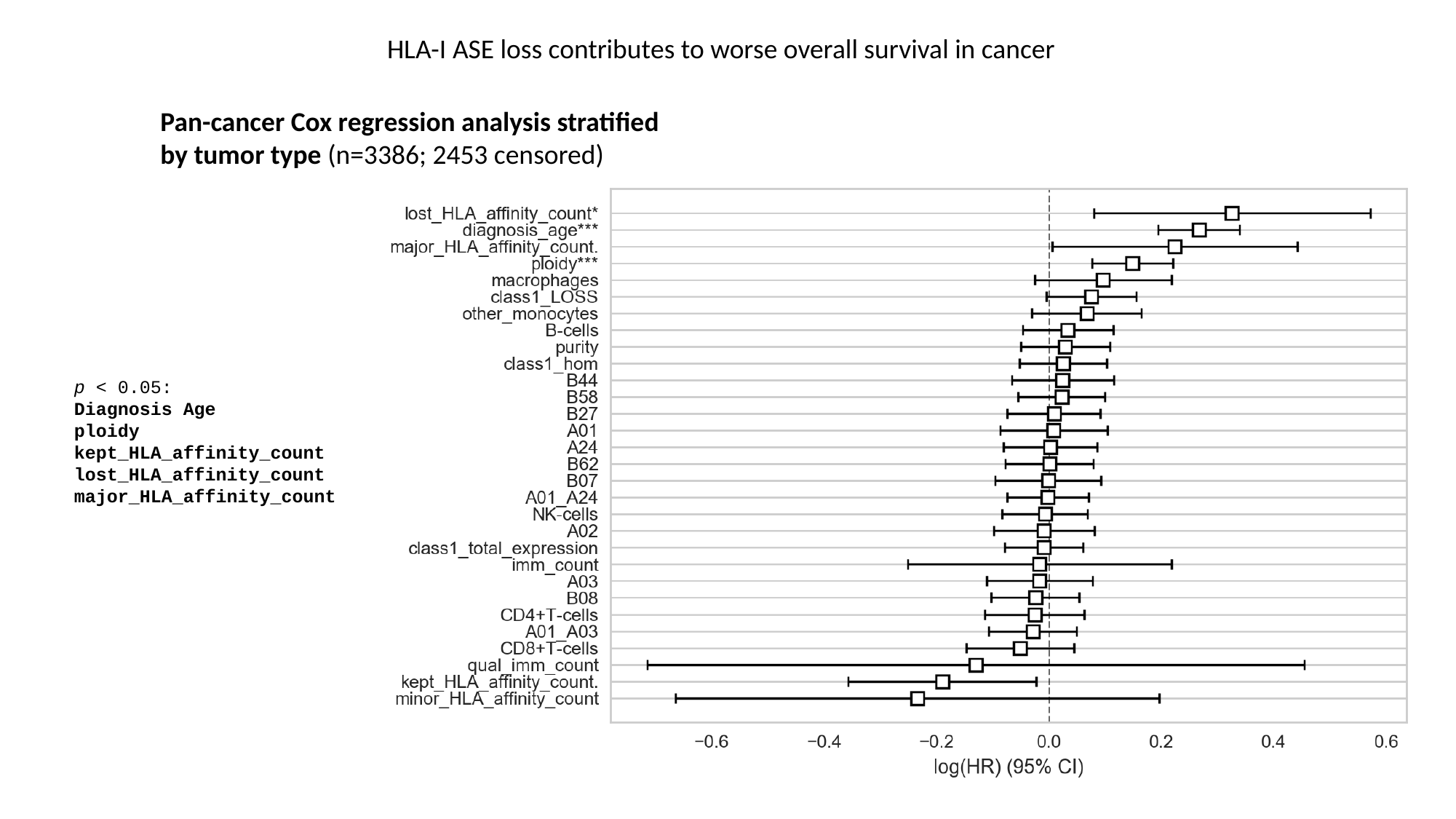

HLA-I ASE loss contributes to worse overall survival in cancer
Pan-cancer Cox regression analysis stratified by tumor type (n=3386; 2453 censored)
p < 0.05:
Diagnosis Age
ploidy
kept_HLA_affinity_count lost_HLA_affinity_count
major_HLA_affinity_count

### Slide 4
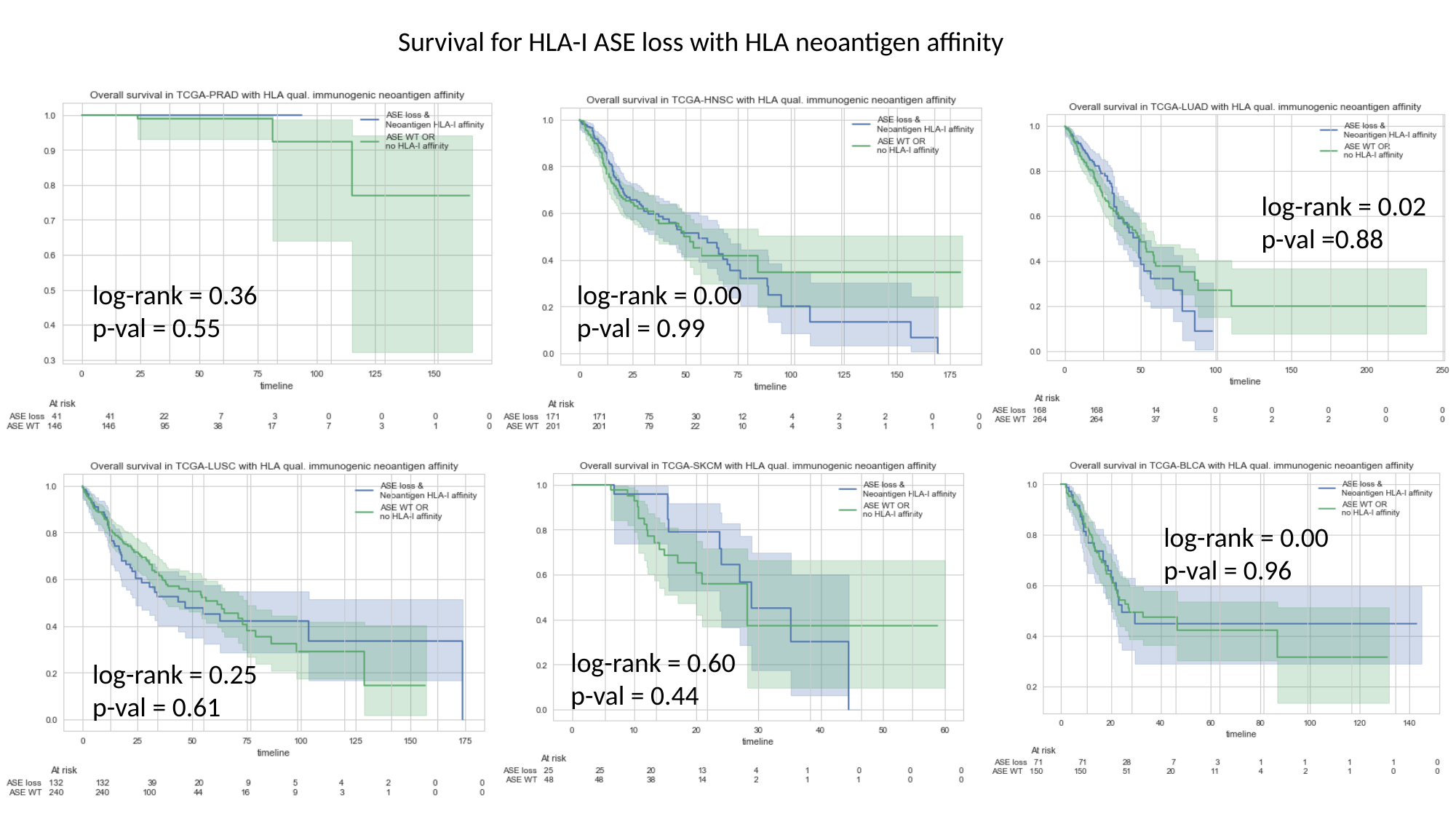

#### Survival for HLA-I ASE loss with HLA neoantigen affinity
log-rank = 0.02
p-val =0.88
log-rank = 0.00
p-val = 0.99
log-rank = 0.36
p-val = 0.55
log-rank = 0.00
p-val = 0.96
log-rank = 0.60
p-val = 0.44
log-rank = 0.25
p-val = 0.61

### Slide 5
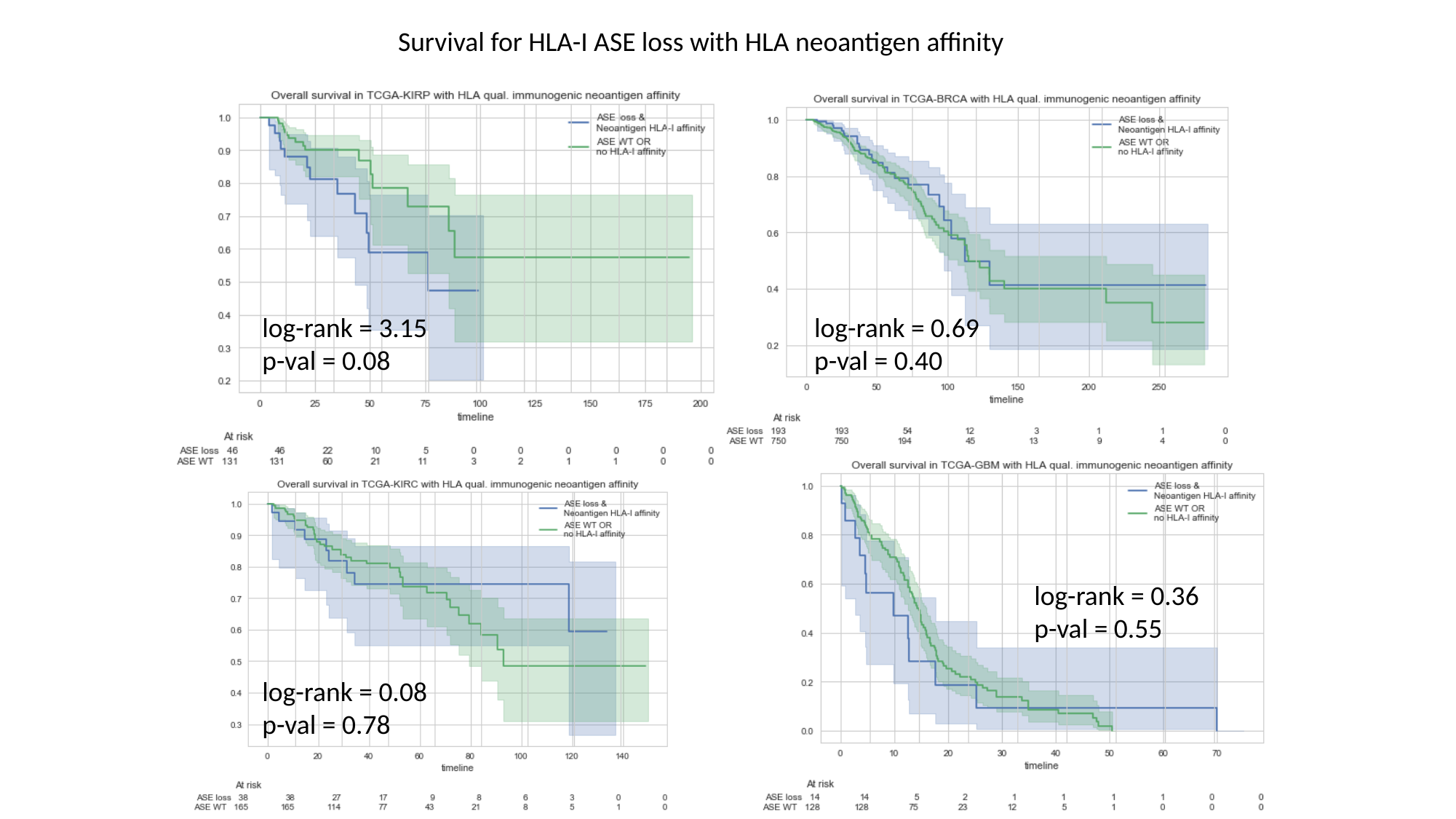

#### Survival for HLA-I ASE loss with HLA neoantigen affinity
log-rank = 0.69
p-val = 0.40
log-rank = 3.15
p-val = 0.08
log-rank = 0.36
p-val = 0.55
log-rank = 0.08
p-val = 0.78

### Slide 6
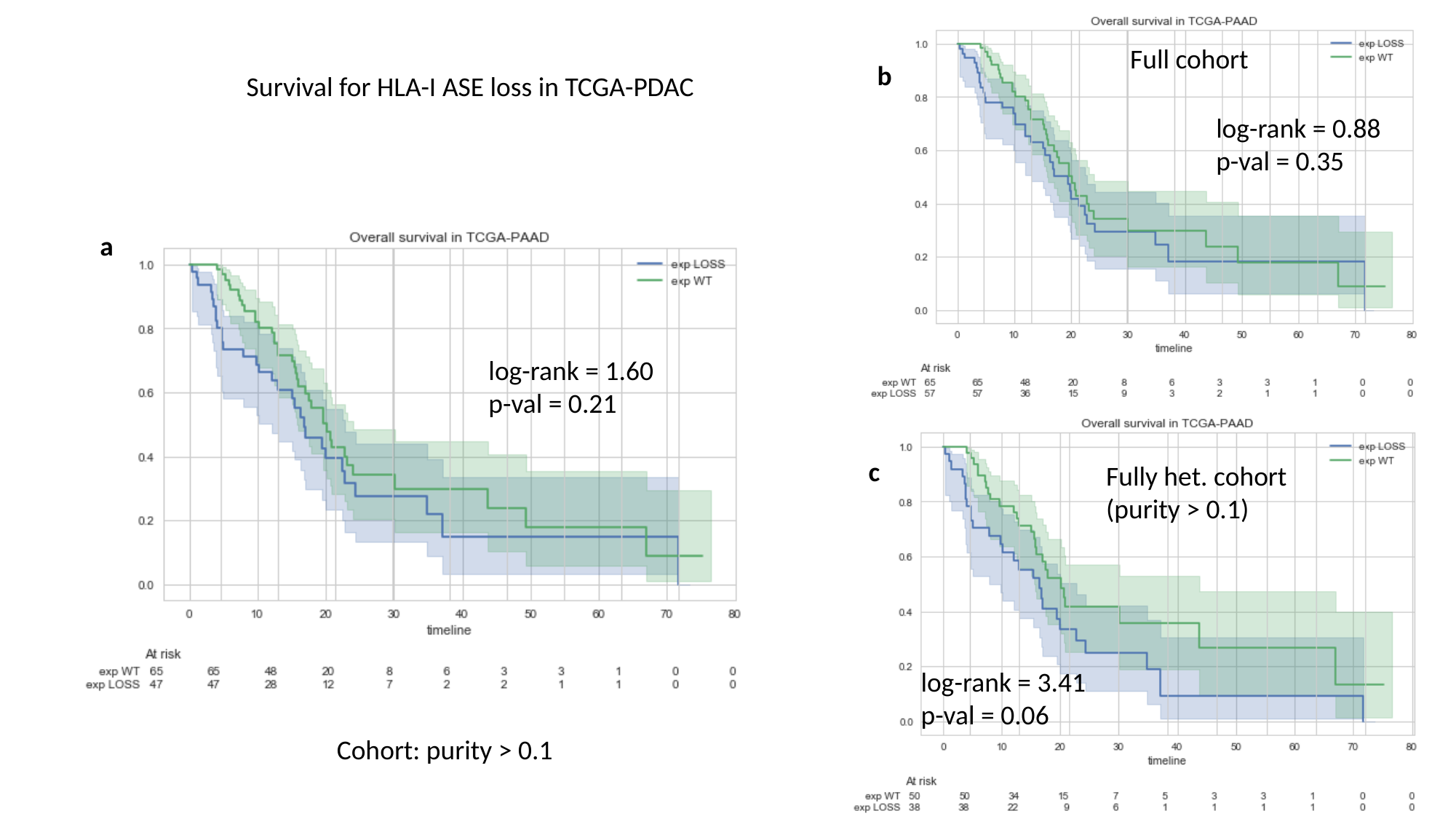

Full cohort
#### Survival for HLA-I ASE loss in TCGA-PDAC
b
log-rank = 0.88
p-val = 0.35
a
log-rank = 1.60
p-val = 0.21
c
Fully het. cohort
(purity > 0.1)
log-rank = 3.41
p-val = 0.06
Cohort: purity > 0.1

### Slide 7
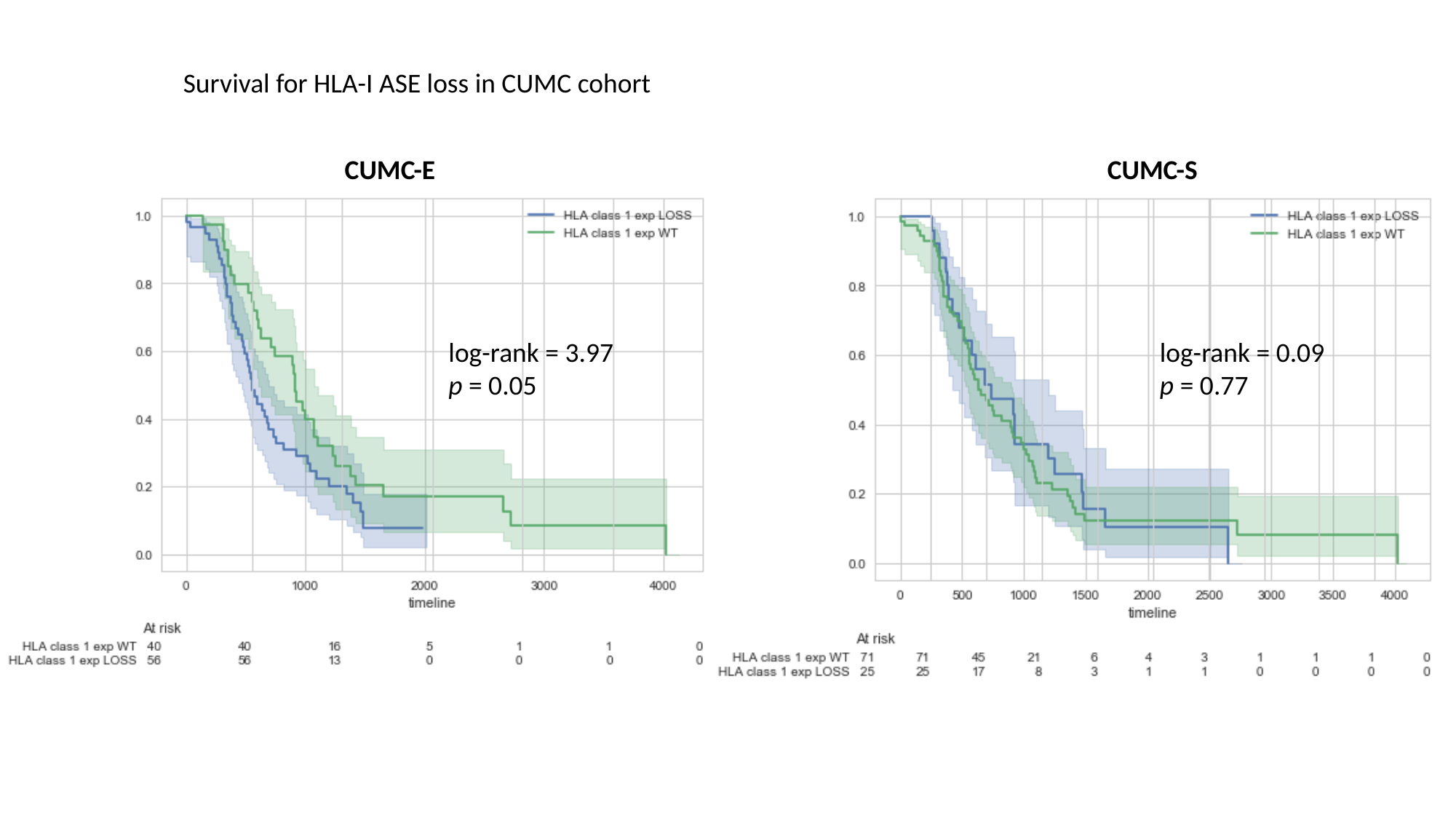

#### Survival for HLA-I ASE loss in CUMC cohort
CUMC-E
CUMC-S
log-rank = 0.09
p = 0.77
log-rank = 3.97
p = 0.05

### Slide 8
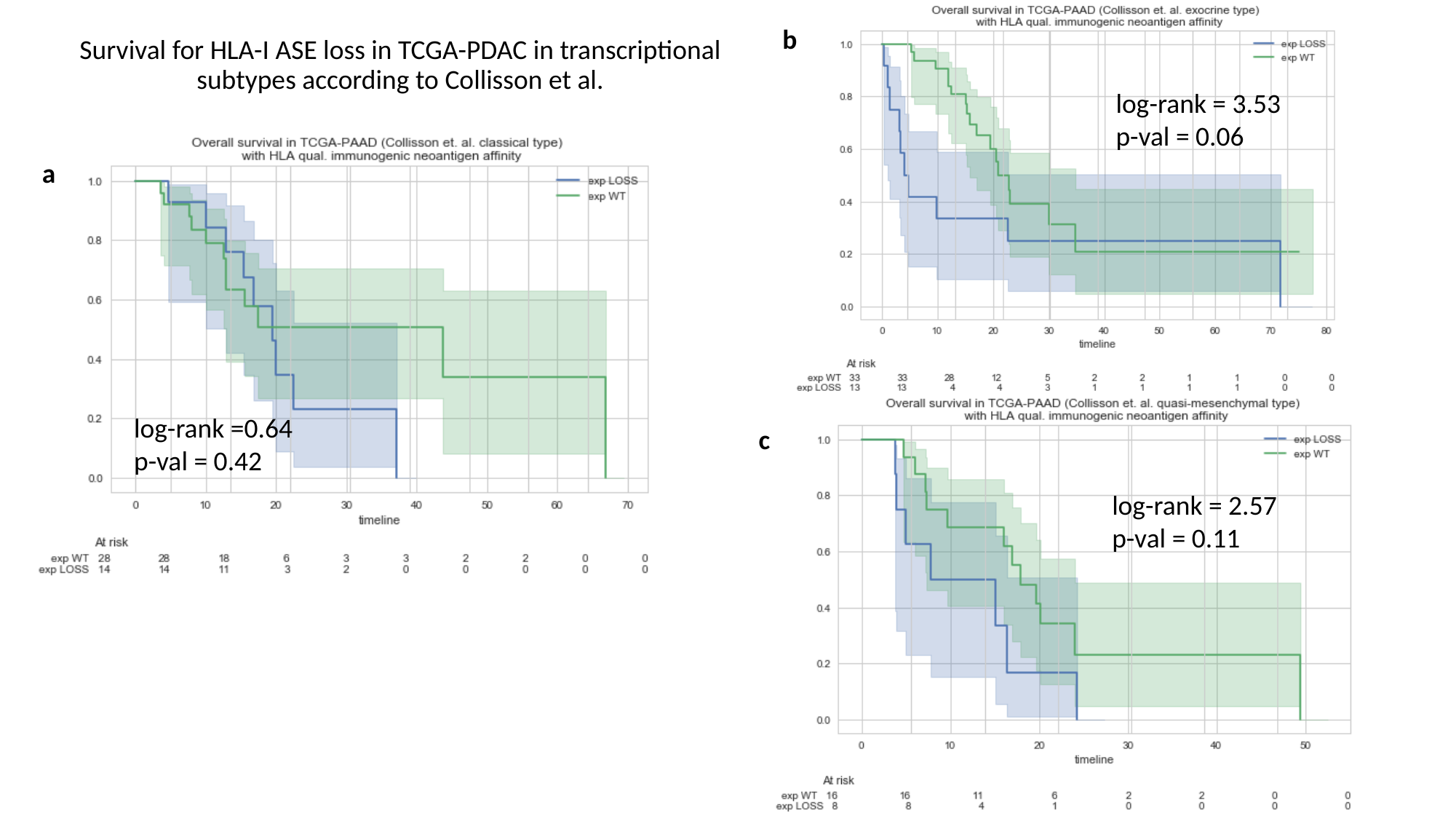

b
#### Survival for HLA-I ASE loss in TCGA-PDAC in transcriptional subtypes according to Collisson et al.
log-rank = 3.53
p-val = 0.06
a
log-rank =0.64
p-val = 0.42
c
log-rank = 2.57
p-val = 0.11

### Slide 9
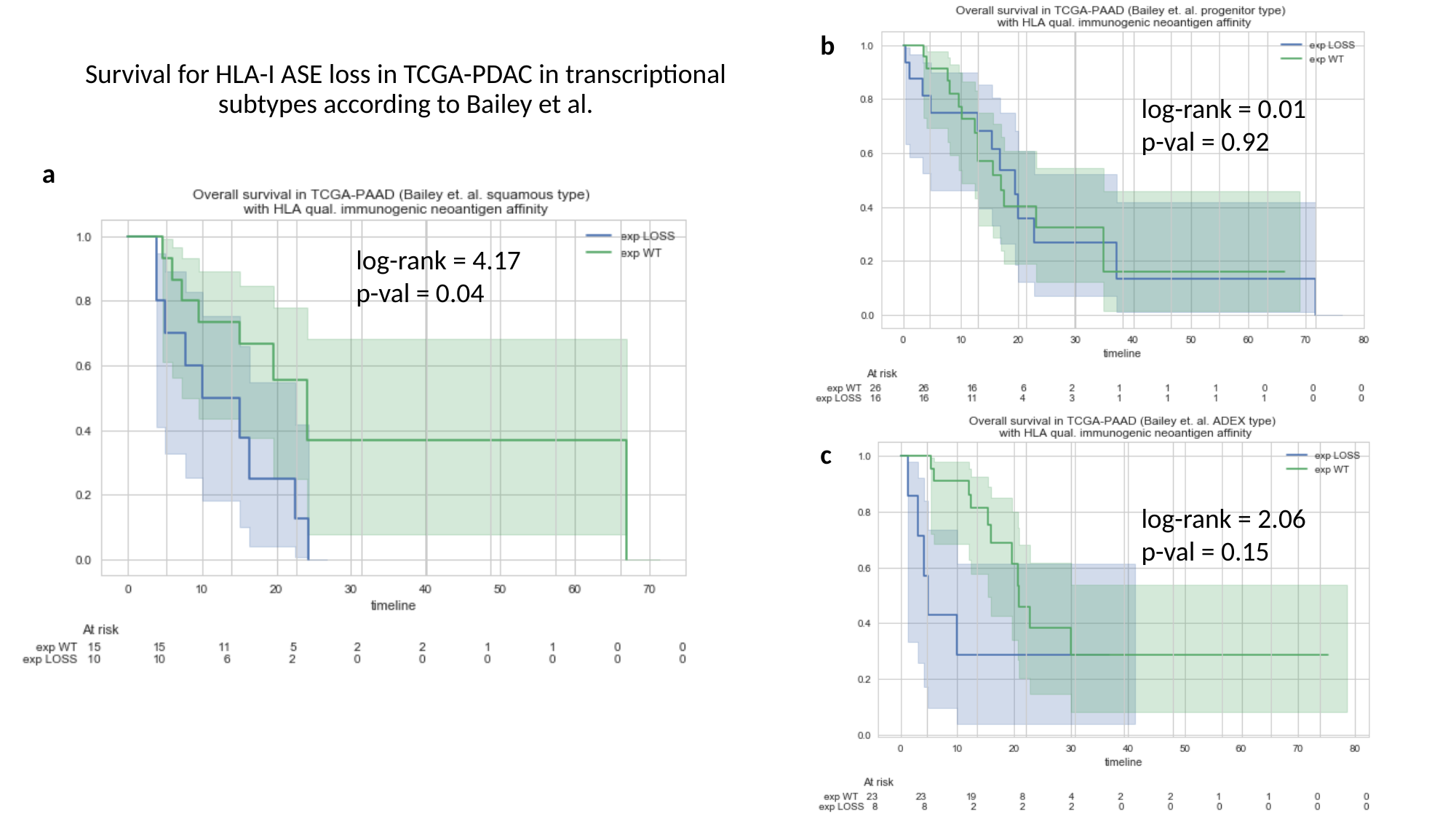

b
#### Survival for HLA-I ASE loss in TCGA-PDAC in transcriptional subtypes according to Bailey et al.
log-rank = 0.01
p-val = 0.92
a
log-rank = 4.17
p-val = 0.04
c
log-rank = 2.06
p-val = 0.15

### Slide 10
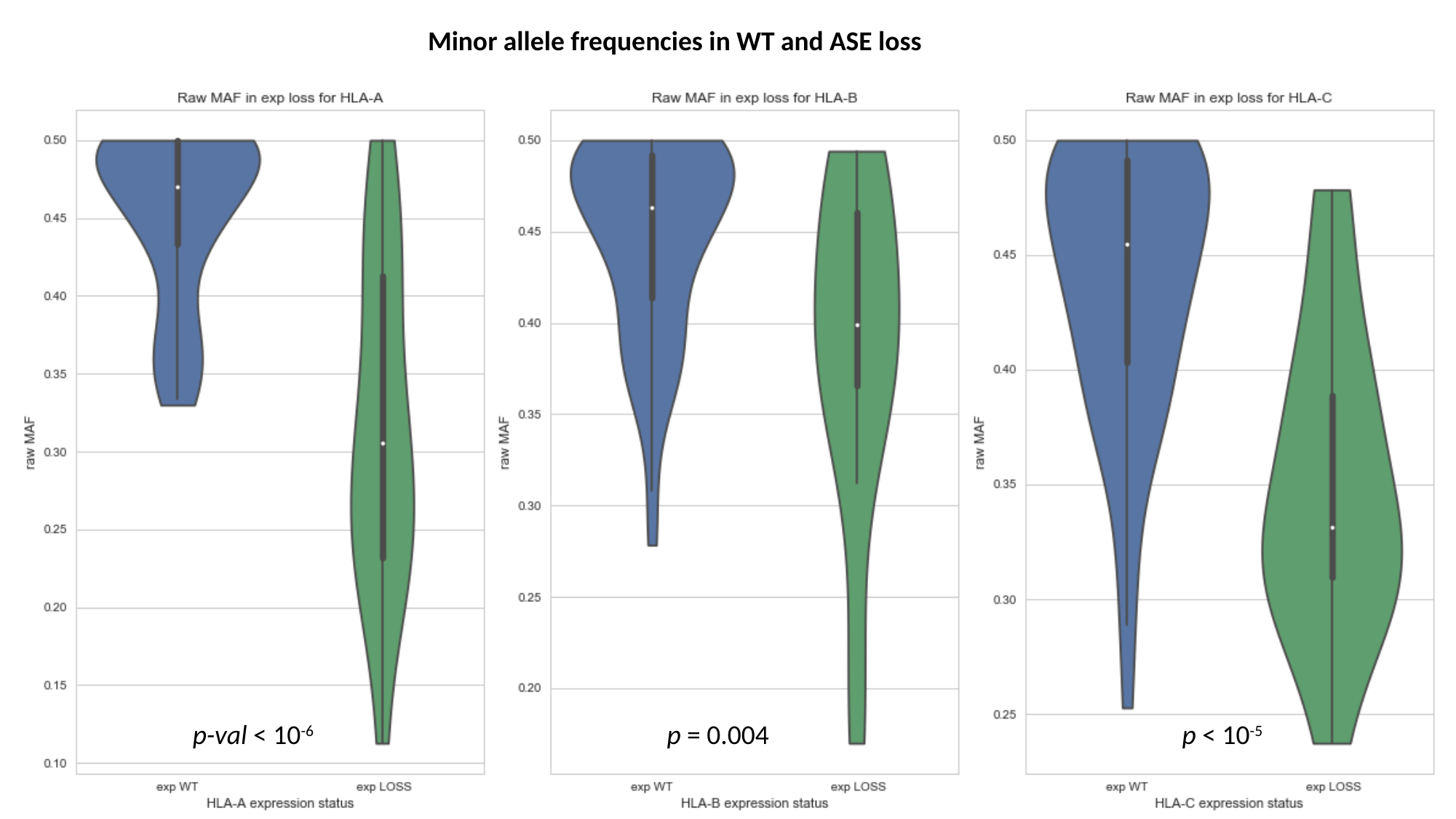

Minor allele frequencies in WT and ASE loss
p-val < 10^-6^
p = 0.004
p < 10^-5^

### Slide 11
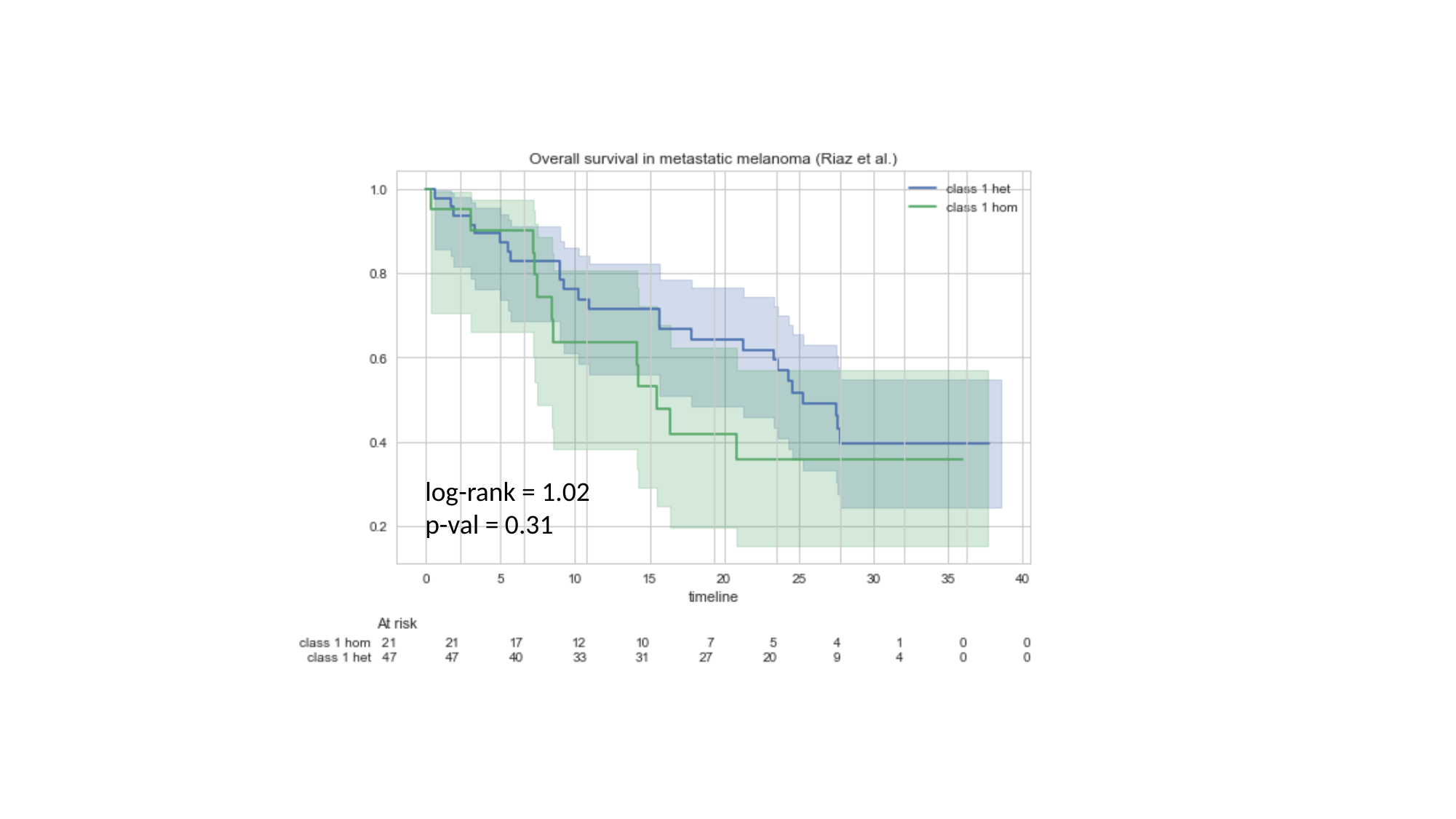

log-rank = 1.02 p-val = 0.31

### Slide 12
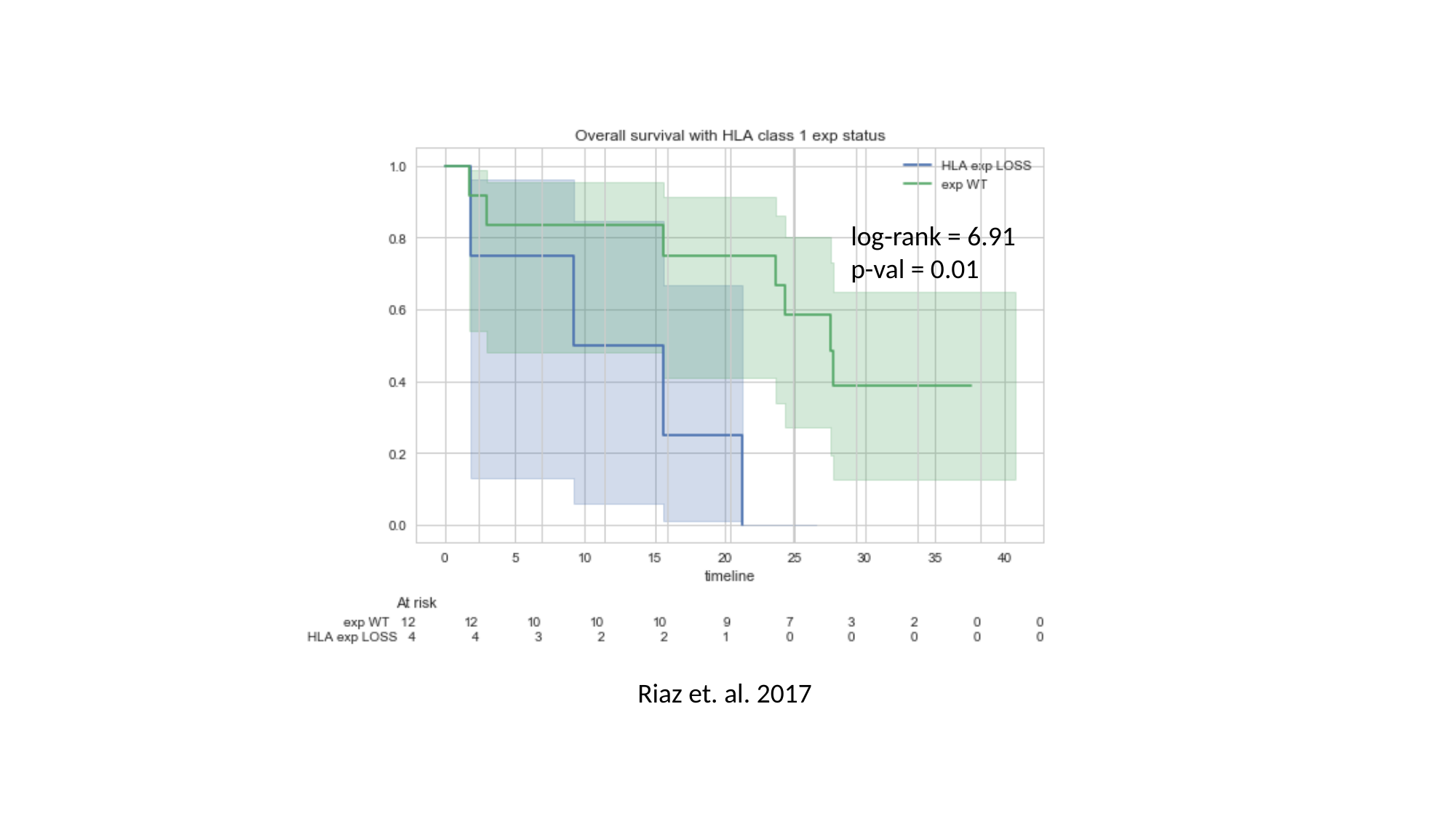

log-rank = 6.91
p-val = 0.01
Riaz et. al. 2017

### Slide 13
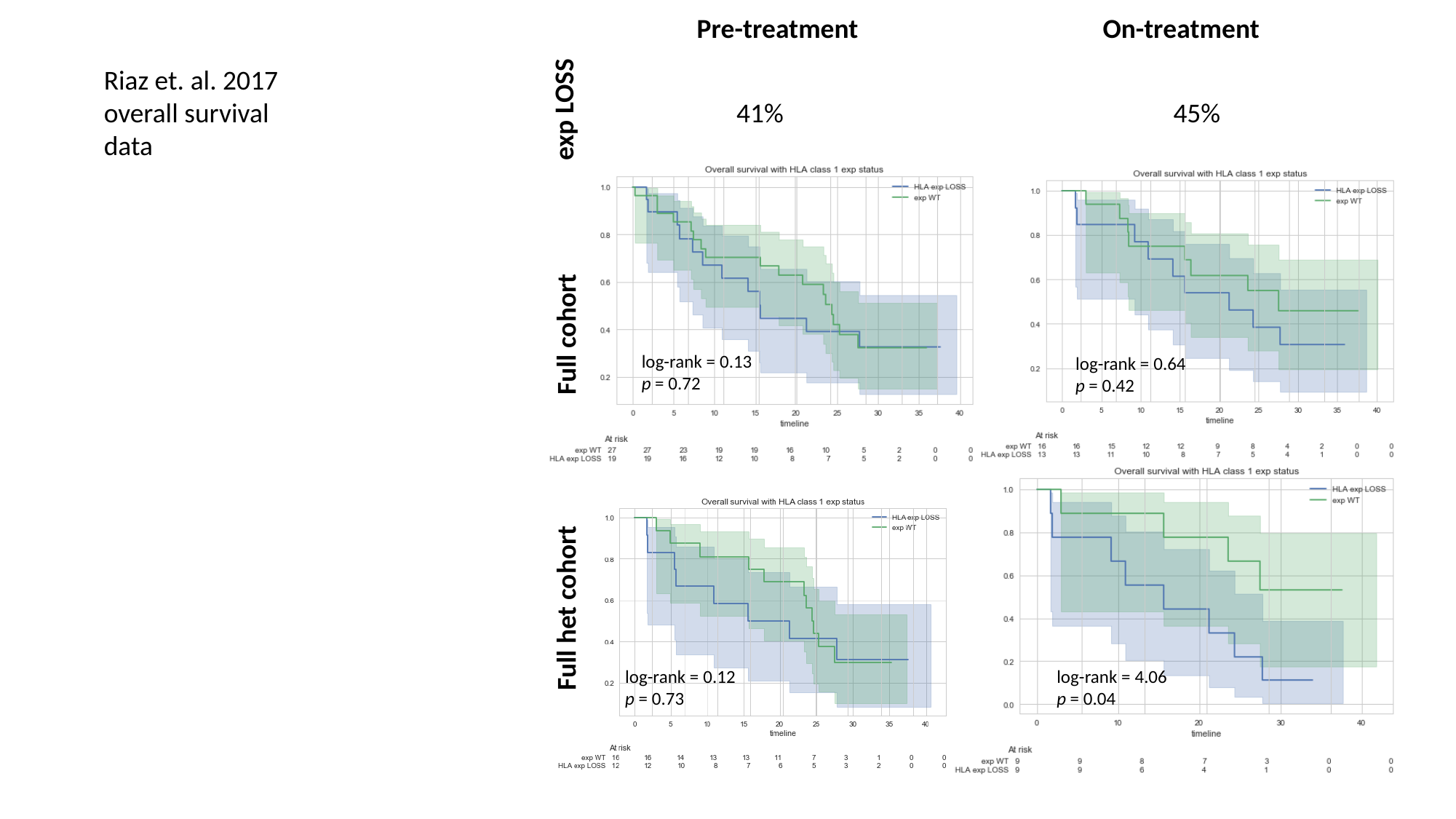

Pre-treatment
On-treatment
Riaz et. al. 2017 overall survival data
exp LOSS
41%
45%
Full cohort
log-rank = 0.13
p = 0.72
log-rank = 0.64
p = 0.42
Full het cohort
log-rank = 0.12
p = 0.73
log-rank = 4.06 p = 0.04

### Slide 14
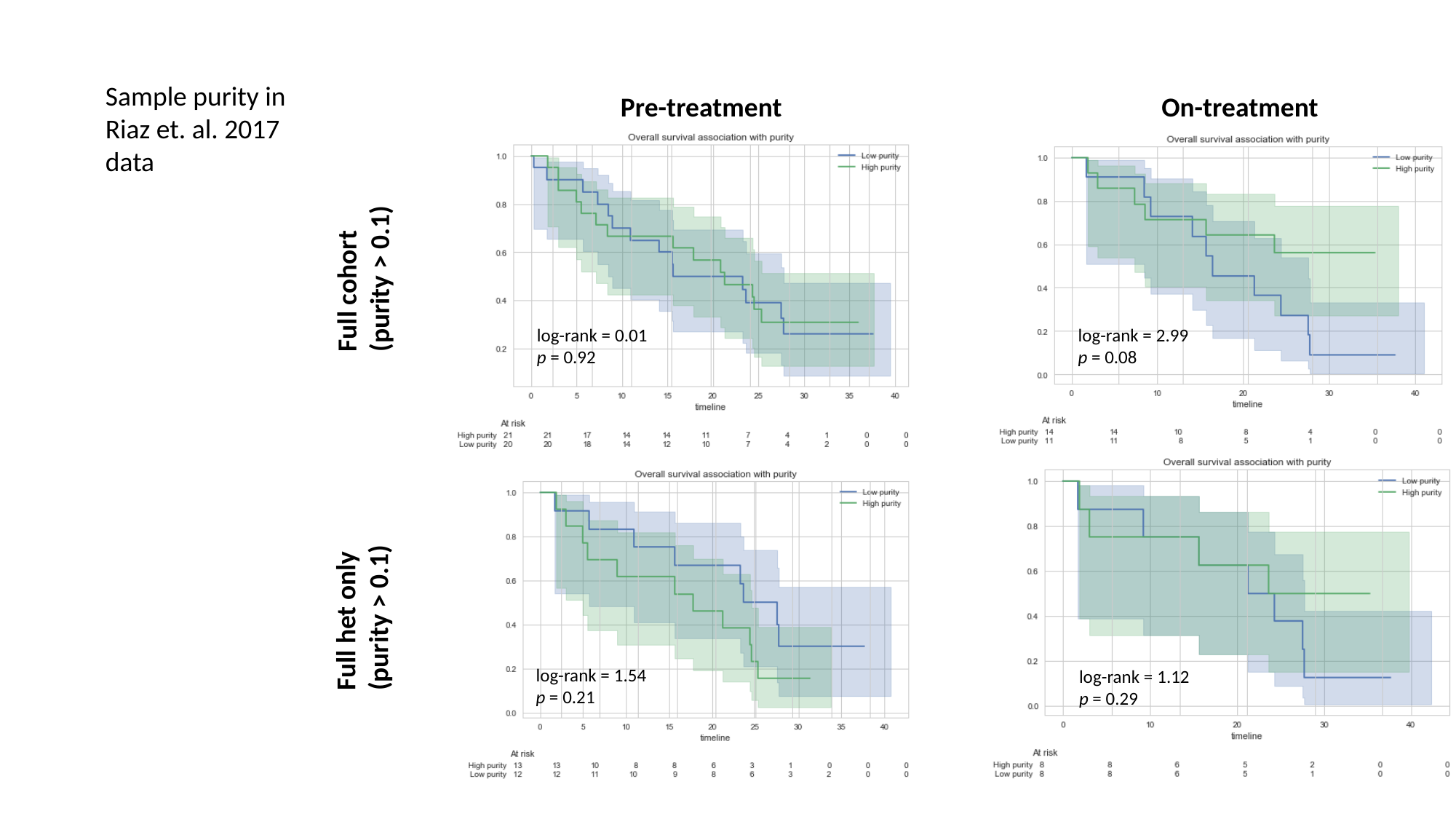

Sample purity in Riaz et. al. 2017 data
Pre-treatment
On-treatment
Full cohort (purity > 0.1)
log-rank = 0.01
p = 0.92
log-rank = 2.99
p = 0.08
Full het only (purity > 0.1)
log-rank = 1.54
p = 0.21
log-rank = 1.12
p = 0.29

### Slide 15
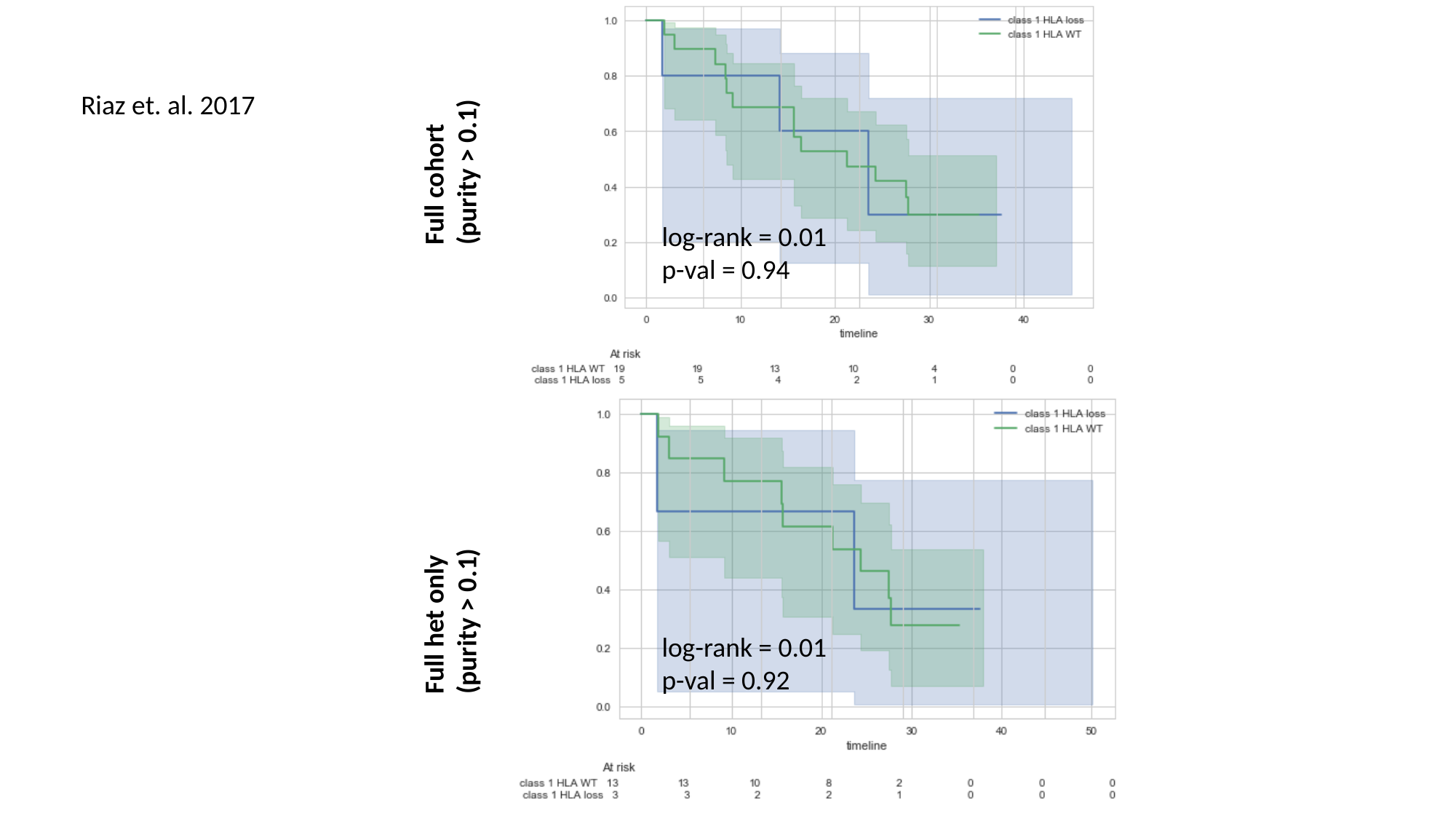

Riaz et. al. 2017
Full cohort (purity > 0.1)
log-rank = 0.01
p-val = 0.94
Full het only (purity > 0.1)
log-rank = 0.01
p-val = 0.92
